# Effect of Continuum of Maternal Healthcare on Neonatal Mortality in Sub-Saharan Africa: A Pooled DHS-8 Analysis

**DOI:** 10.64898/2026.08.18.26360750

**Authors:** Samba Camara, Duah Dwomoh, Prudence Tettey, Amadou Barrow

## Abstract

**Background:** Neonatal mortality remains disproportionately high in sub-Saharan Africa (SSA), accounting for nearly half of all deaths in children under five. Although antenatal care, skilled birth attendance, and postnatal care are individually associated with improved newborn survival, few studies have examined whether their integrated receipt as a continuum of care (CoC) confers additional protection against neonatal death, particularly after accounting for sociodemographic confounding and heterogeneity across country contexts.

**Methods:** A pooled cross-sectional analysis was conducted using DHS-8 Births Recode files from five countries: Nigeria (2024), Mali (2023–2024), Congo DRC (2023–2024), Kenya (2022), and Lesotho (2023–2024). The analytical sample comprised 37,351 births within the 36-month postnatal care reference window. Complete CoC was defined as receipt of adequate antenatal care (≥4 visits with first-trimester initiation), skilled birth attendance, and postnatal care within 48 hours for the mother or newborn. Neonatal mortality was defined as death within 27 days of birth. Survey-weighted logistic and log-Poisson regression models estimated adjusted odds ratios (aOR) and adjusted prevalence ratios (aPR). G-computation quantified the population attributable fraction (PAF). Country-specific heterogeneity was examined through random-effects meta-analysis (DerSimonian-Laird method) and a two-level multilevel logistic regression model.

**Results:** The overall neonatal mortality rate was 29.3 per 1,000 live births (95% CI: 27.6–31.0). Complete CoC prevalence was 19.2% (95% CI: 18.5–19.9%), ranging from 7.8% in Congo DRC to 47.8% in Lesotho. In unadjusted analysis, complete CoC was associated with a 24% reduction in neonatal death odds (cOR: 0.764, 95% CI: 0.583–1.000, p = 0.050). After adjustment for wealth, education, residence, parity, maternal age, child sex, and country, the association was substantially attenuated and non-significant (aOR: 0.961, 95% CI: 0.717–1.289; aPR: 0.962, 95% CI: 0.722–1.282). The PAF under universal complete CoC was 3.2%. The pooled meta-analytic estimate was aOR 0.718 (95% CI: 0.447–1.152), with moderate heterogeneity (I² = 38.9%; τ² = 0.089). Country-specific estimates ranged from aOR 0.455 (95% CI: 0.256–0.810) in Kenya to 1.447 (95% CI: 0.496–4.220) in Lesotho.

**Conclusion:** Complete continuity of maternal healthcare was not independently associated with reduced neonatal mortality after full adjustment, suggesting that the unadjusted benefit was attributable to sociodemographic selection rather than a direct causal pathway. These findings underscore the insufficiency of service utilisation metrics in isolation and highlight the need to address the structural and contextual determinants that simultaneously constrain both care access and neonatal survival. Country-level heterogeneity in the CoC-mortality relationship points to the importance of tailored, context-specific interventions.

## Introduction

Neonatal mortality, defined as death within the first 27 days of life, remains one of the most persistent and inequitably distributed public health challenges of our era. In 2022, an estimated 2.3 million neonates died worldwide, of whom approximately 43% were born in sub-Saharan Africa (SSA), a region that carries a disproportionate share of global neonatal deaths relative to its birth volume [1, 2]. The neonatal mortality rate (NMR) in SSA averaged 27 deaths per 1,000 live births in 2022, compared with 3 per 1,000 in high-income regions, a disparity that has narrowed only modestly over the past two decades [1]. The majority of neonatal deaths in SSA occur within the first seven days of life and are attributable to preventable conditions: birth asphyxia, infection, preterm birth complications, and hypothermia [3, 4]. These are conditions for which timely and skilled obstetric and newborn care represents the most evidence-based intervention available. Achieving Sustainable Development Goal 3.2 which targets a neonatal mortality rate no higher than 12 per 1,000 live births in every country by 2030 is therefore contingent on strengthening maternal and newborn health service delivery across the SSA region [5].

The three principal components of maternal healthcare - antenatal care, skilled birth attendance, and postnatal care - have each been linked to reduced maternal and neonatal mortality in low-income settings [6–8]. Antenatal care enables early identification of obstetric complications, ensures tetanus immunisation and nutrition supplementation, and provides birth preparedness and health education [9]. The World Health Organization recommends eight antenatal contacts minimum, with the first in the first trimester [10]. Skilled birth attendance, defined as delivery by a trained health professional with competency and equipment to manage normal deliveries and complications, is recognized as the most effective intervention to reduce intrapartum and early neonatal mortality [11]. Postnatal care within 48 hours after delivery is critical for newborn survival: most neonatal infections and respiratory complications manifest in this window, and thermal care, breastfeeding support, and resuscitation readiness are most effective immediately after birth [12, 13].

Despite established effectiveness of individual components, coverage data from SSA show fragmented service use: women often initiate antenatal care without completing recommended visits, deliver with skilled attendants without postnatal checks, or attend antenatal care late [14, 15]. This fragmentation led to the continuum of care framework, introduced by Kerber and colleagues [16] and adopted by WHO and UNICEF as a strategy for reducing maternal and neonatal deaths. The framework suggests that sustained engagement with healthcare from pregnancy through postpartum period provides greater survival benefit than isolated contacts by addressing risk across the perinatal period. However, empirical evidence for this integrated hypothesis remains limited in SSA. Most analyses have examined individual components or pairwise combinations, with few studies modeling all three components against defined binary outcomes [15, 17, 18]. Pooled analyses have mainly focused on estimating prevalence and determinants of complete CoC rather than its association with neonatal mortality [17, 18], or been limited to one or two countries [19]. A comprehensive multi-country analysis linking all CoC components to neonatal mortality, with adjustment for confounding, population-level impact estimation, and cross-country heterogeneity quantification, has not been conducted using DHS Phase 8 data.

DHS Phase 8 surveys, conducted between 2019 and 2025, introduced substantive revisions to the measurement of postnatal care that are particularly relevant for continuum of care research. The reference period for postnatal care questions was reduced from six to three years preceding the survey, and a new Pregnancy and Postnatal Care Recode file was introduced, enabling more precise measurement of the timing, provider type, and content of postpartum care for both mothers and newborns [20]. These methodological improvements address longstanding concerns about recall bias in retrospective reports of early postnatal care and permit more valid comparisons of postnatal care coverage across countries. However, they also introduce new analytical challenges, including the need to restrict the continuum of care analysis to the shorter postnatal reference window and the need to account for the revised timing variable coding schema. To date, no pooled analysis of CoC and neonatal mortality has been conducted using DHS-8 data, and the implications of the revised measurement framework for CoC estimates and their association with neonatal outcomes have not been examined. Given the heterogeneity in healthcare system capacity, policy environments, and care-seeking behaviour across SSA, understanding how the CoC-mortality relationship varies between country contexts is equally important for informing national and regional programme design.

The present study addresses this central research question: *Does receiving a complete continuum of maternal healthcare - including adequate antenatal care, skilled birth attendance, and postnatal care within 48 hours - independently reduce neonatal mortality risk in selected sub-Saharan Africa after accounting for sociodemographic determinants?* The primary hypothesis was that complete CoC would significantly reduce neonatal mortality in adjusted analyses, consistent with biological plausibility and evidence from component studies. A secondary hypothesis was that this association would vary across countries, reflecting differences in healthcare delivery quality and effectiveness in SSA. The study had four objectives: (1) to estimate complete CoC prevalence and characterize sociodemographic differentials; (2) to estimate the adjusted association between complete CoC and neonatal mortality; (3) to quantify the population-level impact through counterfactual analysis and population attributable fraction; and (4) to examine cross-country heterogeneity through meta-analysis and multilevel modelling. The analysis uses DHS-8 surveys from Nigeria, Mali, Congo DRC, Kenya, and Lesotho, representing four SSA sub-regions and accounting for over 70 million annual live births.

## Methods

### Study design and data source

This study used a pooled cross-sectional design based on nationally representative Demographic and Health Surveys (DHS) from five sub-Saharan African countries. The DHS programme conducts standardised, nationally representative household surveys in low- and middle-income countries using a stratified two-stage cluster sampling design; methodological details are available in published documentation [21, 22]. Data were obtained from five Phase 8 (DHS-8) surveys: Nigeria 2024, Mali 2023-2024, Congo DRC 2023-2024, Kenya 2022, and Lesotho 2023-2024. DHS-8 represents a methodological update from earlier phases, including the replacement of a full birth history with a full pregnancy history (facilitating enhanced postnatal care data collection), a revised reference period for postnatal care questions (three rather than six years preceding the survey), and the introduction of a dedicated Pregnancy and Postnatal Care Recode (NR) file. All datasets were accessed through a registered DHS Programme account under standard data use agreements.

### Study population and sample

The analytic unit was the live birth. The primary study population comprised all live births occurring within five years preceding each respective survey, as recorded in the Births Recode (BR) file, which contains complete birth histories for all interviewed women aged 15 to 49 years. For the continuum of care analysis, a further restriction was applied: only births within 36 months preceding the survey were included, to align with the postnatal care reference window in DHS-8 [20, 21]. Singleton and multiple births were both included; a sensitivity analysis restricted to singletons (b0 = 0 in DHS notation) was conducted to assess robustness of estimates to multiple gestation.

### Outcome variable: neonatal mortality

The primary outcome was neonatal mortality, defined as death occurring within the first 28 completed days of life, consistent with the World Health Organization (WHO) and DHS standard definition [22, 23]. Neonatal death was identified using a combination of DHS birth history variables: survival status (b5 = 0 indicating deceased), age at death in months (b7 = 0 indicating death in month zero, corresponding to the first 28 days), and age at death in days coded at the precise level (b6 in the range 100 to 127, corresponding to days 0 to 27). Early neonatal death (days 0 to 6) and late neonatal death (days 7 to 27) were also generated as supplementary outcomes.

### Exposure variable: Continuum of maternal healthcare

The primary exposure was the continuum of care (CoC) composite indicator, operationalised as the simultaneous receipt of three components aligned with WHO recommendations for maternal and newborn healthcare [23].

### Component 1: Adequate antenatal care

Adequate ANC was defined as receipt of four or more ANC visits (m14 ≥ 4) with initiation of the first visit during the first trimester (≤3 months of gestation, m13 ≤ 3). This combined definition was adopted based on recent pooled SSA DHS literature demonstrating that both visit frequency and early initiation independently predict neonatal outcomes [17, 18]. Two supplementary definitions were also constructed: ANC4+ alone (without timing requirement, for sensitivity analysis SA1) and ANC8+ as per the WHO 2016 ANC model, which recommends a minimum of eight contacts [23] (sensitivity analysis SA2).

### Component 2: Skilled Birth Attendance

Skilled birth attendance (SBA) was defined as the delivery being assisted by a doctor (m3a = 1), nurse or midwife (m3b = 1), or auxiliary nurse or midwife (m3c = 1), in accordance with the WHO definition of skilled health personnel [24].

### Component 3: Postnatal care within 48 hours

PNC within two days was coded using the DHS-8 specific timing schema for the m-series variables [20, 22]. For the mother, PNC was considered received within two days if: (i) a check before facility discharge was conducted by a health provider (m62 = 1 and m64 within the range 10 to 29), with timing in the range 100 to 201 (hours 0 to 71, or 1 to 2 days), including codes 198 to 199 (hours but value missing or unknown, which DHS codes as equivalent to within two days); or (ii) a check after discharge or at home was conducted by a health provider (m66 = 1 and m68 in range 10 to 29) with the same timing criteria. The same logic was applied for newborn PNC using m74/m75/m76 (facility discharge check) and m70/m71/m72 (subsequent check), as specified in the DHS Guide to Statistics and the DHS Programme code library for DHS-8 surveys [20, 22]. The primary combined PNC component was defined as either the mother or the newborn (or both) receiving PNC within two days. A strict definition requiring both mother and newborn to receive PNC was used in sensitivity analysis SA4.

The binary complete CoC variable was coded as 1 if a woman received all three components, and 0 otherwise. A continuous CoC score (0 to 3) representing the number of components received was also constructed for dose-response analysis.

### Covariates

Covariates were selected a priori based on a conceptual framework adapted from Asratie et al. [17] and the established literature on determinants of neonatal mortality in SSA [3, 4]. Individual-level covariates included: household wealth quintile (v190; 1 = poorest, 5 = richest), maternal educational attainment (v106; none, primary, secondary, higher), place of residence (v025; urban, rural), parity category (derived from birth order bord: primipara, 2nd to 3rd, 4th to 5th, 6th or higher), maternal age group (v013; 15 to 19, 20 to 24, 25 to 29, 30 to 34, 35 to 39, 40 to 44, 45 to 49 years), sex of the child (b4), and preceding birth interval (b11). Country was included as a fixed effect in all pooled models to account for unmeasured between-country confounding.

### Statistical analysis

#### Survey design and weighting

All analyses accounted for the complex two-stage stratified cluster sampling design of the DHS using Stata’s *svyset* command [25]. The primary sampling unit was the DHS cluster (v021), the stratification variable was v022, and individual sample weights were applied (v005 divided by 1,000,000). For the pooled analysis, weights were normalised within each country by dividing the individual weight by the country-specific mean weight, so that each country contributed proportionally to its sample size rather than its population size [17, 26]. Unique cluster and stratum identifiers were constructed to prevent ID collisions across countries in the pooled dataset. The variance estimator was linearised (Taylor series), with the *singleunit(centered)* option applied for strata with a single sampling unit [25].

### Objective 1: CoC prevalence and sociodemographic correlates

Survey-weighted proportions with 95% confidence intervals (CIs) were estimated for each CoC component and the composite CoC indicator, overall and by country, using the *svy: proportion* and *svy: mean* commands restricted to the CoC analysis subpopulation (in_coc = 1) [25]. Design effects (DEFFs) were reported to characterise the efficiency loss attributable to cluster sampling. Bivariate cross-tabulations of CoC status by each sociodemographic covariate were examined using the *svy: tab* command, with design-adjusted Pearson F-statistics used for significance testing [25].

### Objective 2: Association between CoC and neonatal mortality

The association between complete CoC and neonatal mortality was estimated using two complementary regression approaches [27]. First, survey-weighted logistic regression (*svy: logistic*) was used to estimate unadjusted (crude) odds ratios (cOR) and adjusted odds ratios (aOR) with 95% CIs. Second, survey-weighted log-binomial regression via the Poisson approximation (*svy: poisson, irr*) was used to estimate adjusted prevalence ratios (aPR), which are more interpretable than odds ratios when the outcome is common [28, 29]. Poisson models with robust variance were preferred over binomial models to avoid convergence issues. Binary CoC and the four-category CoC score (0 to 3) were fitted in separate models; the zero-component category served as the reference group for the dose-response analysis. A formal test for linear trend across the CoC score was conducted by treating the score as a continuous variable in the logistic model and applying a Wald test [30]. Both logistic and Poisson models were adjusted simultaneously for all covariates listed above, with country included as a categorical fixed effect.

### Objective 3: Counterfactual analysis

G-computation (also termed marginal standardisation or the method of recycled predictions) was applied to quantify the population-level impact of complete CoC under counterfactual scenarios [31]. From the fitted logistic regression model, individual predicted probabilities of neonatal death were generated under:

(i) observed CoC status; (ii) the counterfactual scenario in which all women receive complete CoC; and (iii) the counterfactual scenario in which no woman receives complete CoC. Survey-weighted means of these predicted probabilities were computed over the analytic subpopulation. The Population Attributable Fraction (PAF) was calculated as (P*observed* – P*counterfactual*) / P*observed* × 100, representing the proportion of neonatal deaths preventable under universal complete CoC [31, 32]. The absolute risk reduction (ARR) and number needed to treat (NNT) were also computed. Survey-weighted average marginal effects (AMEs) of complete CoC were estimated using the *margins* command [25].

### Objective 4: Cross-country heterogeneity

Country-specific adjusted effect estimates were obtained from separate survey-weighted logistic regression models fitted within each country, without the country fixed effect. The log(OR) and corresponding standard errors were extracted from the estimated coefficient matrices for the complete CoC indicator and entered into a DerSimonian-Laird (DL) random-effects meta-analysis using the *metan* command in Stata [33, 34]. The DL method was selected as it is robust under a limited number of studies and does not rely on distributional assumptions about between-study heterogeneity [34]. Heterogeneity was quantified using Cochran’s Q test, the I² statistic (proportion of total variation in the effect estimate attributable to between-study heterogeneity), and the between-study variance τ² [35]. A formal interaction test (CoC × country) in the pooled logistic model was used to assess statistical evidence for effect modification by country [36]. Multilevel logistic regression with a random intercept for country was fitted as a supplementary model using *melogit* to partition individual and country-level variance and estimate the intraclass correlation coefficient (ICC) [37]. A random-slope extension was tested against the random-intercept model using a likelihood ratio test.

### Sensitivity analyses

Four pre-specified sensitivity analyses were conducted. SA1 used a simplified CoC definition requiring ANC4+ (without first-trimester timing), SBA, and PNC within two days, to assess whether the timing requirement for ANC drove the primary results. SA2 replaced the ANC4+ component with ANC8+ per the 2016 WHO ANC model [23], to examine results under a more stringent ANC standard. SA3 restricted the analytic sample to singleton births (b0 = 0) to exclude the influence of multiple gestations, which carry substantially elevated neonatal mortality risk [38]. SA4 used a strict PNC definition requiring both the mother and the newborn to receive a postnatal check within two days of birth. Two post-hoc effect modification analyses examined whether the association between CoC and neonatal mortality varied by place of residence (SA5) and by wealth quintile (SA6), using interaction terms and formal Wald tests.

### Missing data

Missing data occurred primarily in the ANC component (15,044 missing within the PNC reference window) and the PNC component (6,532 missing), largely because DHS-8 collects these variables only for births within the three-year reference period while the five-year birth history extends beyond this window. There were no missing values for any sociodemographic covariate. The primary analysis followed a complete-case approach, which is standard in DHS analysis given the survey-weighted design [21]. The implications of missing CoC component data for the external validity of estimates are acknowledged as a limitation.

All analyses were conducted in Stata version 17 (StataCorp LLC, College Station, TX). Survey-weighted analyses used Stata’s *svy* suite of commands. Random-effects meta-analysis was conducted using the *metan* package (version 4.06) [33]. Multilevel models were fitted using *melogit*. Marginal effects and G-computation were implemented using the *margins* command. Graphs were produced using Stata’s *marginsplot* and the *metan* forest plot functionality. Statistical significance was assessed at the conventional two-tailed α = 0.05 threshold throughout.

### Ethical considerations

The DHS Programme datasets are publicly available anonymised secondary data. All surveys were conducted with informed consent of participants, in accordance with the ICF Institutional Review Board and national ethics committees of each country. No additional ethical approval was required for this secondary analysis. Data access was granted through the DHS Programme’s standard user registration process.

## Results

### Characteristics of the study population

A total of 89,013 births occurring within five years preceding the respective surveys were identified across the five countries. After restricting to births within the 36-month postnatal care reference window and applying complete case criteria, the final analytical sample comprised 37,351 births (Nigeria: 9,772; Kenya: 9,916; Congo DRC: 9,717; Mali: 6,520; Lesotho: 1,426). The analytical sample represented 41.9% of all five-year births, with missingness driven primarily by the stricter PNC reference window in DHS-8 (15,044 missing for the ANC component; 6,532 missing for PNC) rather than covariate incompleteness, as no missing data were observed for maternal age, education, wealth, residence, parity, or child sex.

Of the 37,351 births in the analytical sample, 6,536 (17.5%) had mothers who received complete continuity of maternal healthcare, while the remaining 30,815 (82.5%) received incomplete care. Table 1 presents the sociodemographic characteristics of the study population by CoC status. Complete CoC receipt varied substantially across all sociodemographic strata (all p < 0.001). The wealth gradient was pronounced: only 8.5% of births in the poorest quintile were attended by mothers with complete CoC, compared to 38.5% in the richest quintile. Mothers with higher education were nearly four times as likely to receive complete CoC (39.5%) compared to those with no education (9.4%). Urban residence was associated with a substantially higher prevalence of complete CoC (25.2% vs. 13.4% rural). Primiparous women were most likely to receive complete CoC (23.6%), with coverage declining monotonically with increasing birth order to 9.0% among women of sixth or higher birth order.

**Table 1.** Characteristics of the study population stratified by continuum of care status (n = 37,351 births within the 36-month postnatal care reference window).

| Characteristic | Total (n=37,351) | Incomplete CoC (n=30,815) | Complete CoC (n=6,536) | p-value<sup>a</sup> |
| --- | --- | --- | --- | --- |
| **Country** |  |  |  |  |
| Nigeria | 9,772 (26.2) | 8,211 (84.0) | 1,561 (16.0) | <0.001 |
| Mali | 6,520 (17.5) | 4,835 (74.2) | 1,685 (25.8) |  |
| Congo DRC | 9,717 (26.0) | 9,237 (95.1) | 480 (4.9) |  |
| Kenya | 9,916 (26.5) | 7,762 (78.3) | 2,154 (21.7) |  |
| Lesotho | 1,426 (3.8) | 770 (54.0) | 656 (46.0) |  |
| **Wealth quintile** |  |  |  |  |
| Poorest | 9,048 (24.2) | 8,275 (91.5) | 773 (8.5) | <0.001 |
| Poorer | 7,386 (19.8) | 6,557 (88.8) | 829 (11.2) |  |
| Middle | 7,668 (20.5) | 6,537 (85.3) | 1,131 (14.7) |  |
| Richer | 7,437 (19.9) | 5,873 (79.0) | 1,564 (21.0) |  |
| Richest | 5,812 (15.6) | 3,573 (61.5) | 2,239 (38.5) |  |
| **Maternal education** |  |  |  |  |
| No education | 10,299 (27.6) | 9,330 (90.6) | 969 (9.4) | <0.001 |
| Primary | 9,256 (24.8) | 7,948 (85.9) | 1,308 (14.1) |  |
| Secondary | 14,352 (38.4) | 11,453 (79.8) | 2,899 (20.2) |  |
| Higher | 3,444 (9.2) | 2,084 (60.5) | 1,360 (39.5) |  |
| **Place of residence** |  |  |  |  |
| Urban | 12,929 (34.6) | 9,673 (74.8) | 3,256 (25.2) | <0.001 |
| Rural | 24,422 (65.4) | 21,142 (86.6) | 3,280 (13.4) |  |
| **Maternal age group** |  |  |  |  |
| 15–19 years | 2,885 (7.7) | 2,471 (85.6) | 414 (14.4) | <0.001 |
| 20–24 years | 9,379 (25.1) | 7,728 (82.4) | 1,651 (17.6) |  |
| 25–29 years | 9,498 (25.4) | 7,694 (81.0) | 1,804 (19.0) |  |
| 30–34 years | 7,572 (20.3) | 6,166 (81.4) | 1,406 (18.6) |  |
| 35–39 years | 5,325 (14.3) | 4,454 (83.6) | 871 (16.4) |  |
| 40–44 years | 2,214 (5.9) | 1,886 (85.2) | 328 (14.8) |  |
| 45–49 years | 478 (1.3) | 416 (87.0) | 62 (13.0) |  |
| **Parity** |  |  |  |  |
| Primipara (1st birth) | 8,456 (22.6) | 6,460 (76.4) | 1,996 (23.6) | <0.001 |
| 2nd–3rd birth | 13,381 (35.8) | 10,669 (79.7) | 2,712 (20.3) |  |
| 4th–5th birth | 8,221 (22.0) | 7,050 (85.8) | 1,171 (14.2) |  |
| 6th or higher birth | 7,293 (19.5) | 6,636 (90.9) | 657 (9.0) |  |
| **Neonatal death** |  |  |  |  |
| No | 36,622 (98.0) | 30,184 (98.0) | 6,433 (98.4) | 0.050 |
| Yes | 729 (2.0) | 631 (2.0) | 103 (1.6) |  |

**Note.** Values are unweighted observation counts with weighted column percentages in parentheses, except where indicated. ANC = antenatal care; CoC = continuum of care; SBA = skilled birth attendance; PNC = postnatal care. <sup>a</sup> Design-adjusted Pearson chi-square test from complex survey analysis. Percentage distributions are weighted.

### Prevalence of the maternal healthcare continuum and its components

Table 2 presents the weighted pooled and country-specific coverage of each CoC component. Across the five countries, 65.0% (95% CI: 64.1–65.9%) of mothers received four or more antenatal care visits, but only 30.9% (95% CI: 30.1–31.7%) initiated ANC in the first trimester. When combined into the adequate ANC component (four or more visits with first-trimester initiation), coverage fell to 27.3% (95% CI: 26.5– 28.0%). Skilled birth attendance was the highest-coverage component at 80.0% (95% CI: 79.0–81.0%), though this concealed substantial country heterogeneity: coverage ranged from 61.3% in Nigeria to 93.6% in Lesotho. Postnatal care within two days was received by 57.5% (95% CI: 56.4–58.6%) of mothers and 59.5% (95% CI: 58.4–60.5%) of newborns. When combined, 64.8% (95% CI: 63.7–65.8%) had either the mother or the newborn (or both) receive PNC within two days of birth. The overall pooled prevalence of complete CoC was 19.2% (95% CI: 18.5–19.9%). Country-level estimates varied markedly, from 7.8% in Congo DRC to 47.8% in Lesotho. Nigeria (15.0%), Mali (26.4%), and Kenya (25.6%) occupied intermediate positions. Regarding the CoC score distribution, 10.9% of mothers received none of the three components, 25.3% received exactly one, 44.6% received two, and 19.2% received all three (complete CoC).

**Table 2.**
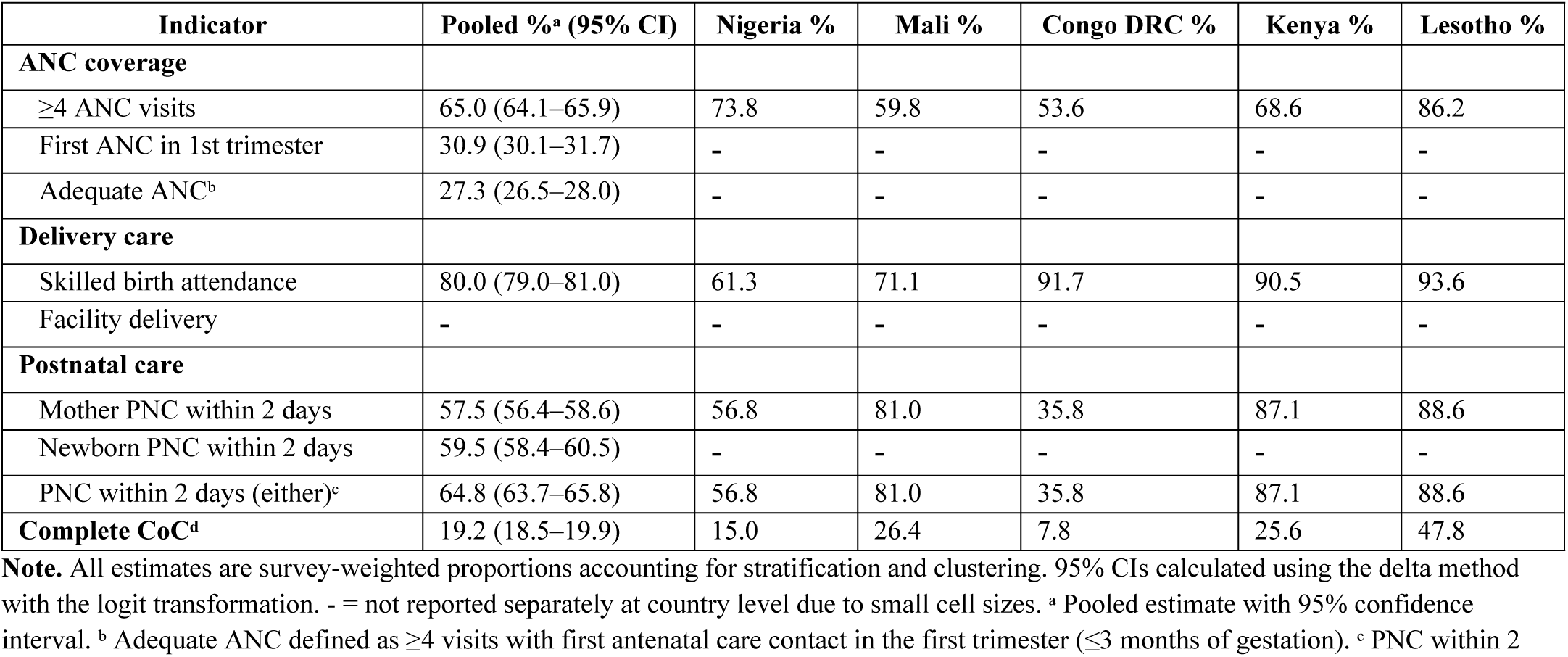
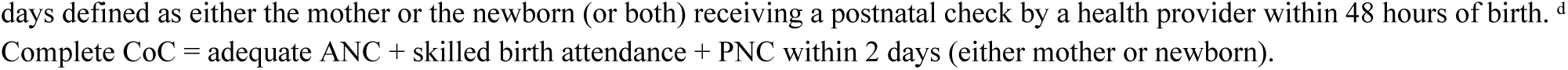
Weighted prevalence of continuum of care components, pooled and by country.

| Indicator | Pooled %<sup>a</sup> (95% CI) | Nigeria % | Mali % | Congo DRC % | Kenya % | Lesotho % |
| --- | --- | --- | --- | --- | --- | --- |
| **ANC coverage** |  |  |  |  |  |  |
| ≥4 ANC visits | 65.0 (64.1–65.9) | 73.8 | 59.8 | 53.6 | 68.6 | 86.2 |
| First ANC in 1st trimester | 30.9 (30.1–31.7) | - | - | - | - | - |
| Adequate ANC<sup>b</sup> | 27.3 (26.5–28.0) | - | - | - | - | - |
| **Delivery care** |  |  |  |  |  |  |
| Skilled birth attendance | 80.0 (79.0–81.0) | 61.3 | 71.1 | 91.7 | 90.5 | 93.6 |
| Facility delivery | - | - | - | - | - | - |
| **Postnatal care** |  |  |  |  |  |  |
| Mother PNC within 2 days | 57.5 (56.4–58.6) | 56.8 | 81.0 | 35.8 | 87.1 | 88.6 |
| Newborn PNC within 2 days | 59.5 (58.4–60.5) | - | - | - | - | - |
| PNC within 2 days (either)<sup>c</sup> | 64.8 (63.7–65.8) | 56.8 | 81.0 | 35.8 | 87.1 | 88.6 |
| **Complete CoC<sup>d</sup>** | 19.2 (18.5–19.9) | 15.0 | 26.4 | 7.8 | 25.6 | 47.8 |

**Note.** All estimates are survey-weighted proportions accounting for stratification and clustering. 95% CIs calculated using the delta method with the logit transformation. - = not reported separately at country level due to small cell sizes. <sup>a</sup> Pooled estimate with 95% confidence interval. <sup>b</sup> Adequate ANC defined as ≥4 visits with first antenatal care contact in the first trimester (≤3 months of gestation). <sup>c</sup> PNC within 2
days defined as either the mother or the newborn (or both) receiving a postnatal check by a health provider within 48 hours of birth. <sup>d</sup> Complete CoC = adequate ANC + skilled birth attendance + PNC within 2 days (either mother or newborn).

### Sociodemographic differentials in continuum of care utilisation

Significant sociodemographic differentials were observed across all strata examined (Table 3). The design-adjusted chi-square tests confirmed statistically significant associations between complete CoC receipt and all covariates: wealth quintile (F[3.77, 16,528] = 310.5, p < 0.001), maternal education (F[2.96, 12,977] = 315.6, p < 0.001), place of residence (F[1, 4,382] = 297.0, p < 0.001), parity (F[2.91, 12,767] = 135.6, p < 0.001), maternal age group (F[5.85, 25,646] = 6.2, p < 0.001), and country (F[3.60, 15,759] = 194.1, p < 0.001). The wealth and education gradients were the most pronounced. Among mothers with secondary education, complete CoC prevalence (20.2%) was approximately double that among mothers with no education (9.4%), and mothers with higher education had the highest coverage at 39.5%. The parity gradient was inversely related: complete CoC coverage was highest among primiparous women (23.6%) and declined to 9.0% among women of sixth or higher parity, suggesting increasing barriers to sustained care engagement with increasing family size.

**Table 3.** Weighted prevalence of complete continuum of care by sociodemographic characteristic across five sub-Saharan African countries (pooled DHS-8 analysis, n = 37,351).

| Characteristic | Total (n = 37,351) | CoC status |  | Design-adjusted F-statistic<sup>a</sup> | p-value |
| --- | --- | --- | --- | --- | --- |
|  |  | Incomplete CoC % (n) | Complete CoC % (n) |  |  |
| **Country** |  |  |  |  |  |
| Nigeria | 9,772 (26.2) | 84.0 (8,211) | 16.0 (1,561) |  |  |
| Mali | 6,520 (17.5) | 74.2 (4,835) | 25.8 (1,685) |  |  |
| Congo DRC | 9,717 (26.0) | 95.1 (9,237) | 4.9 (480) |  |  |
| Kenya | 9,916 (26.5) | 78.3 (7,762) | 21.7 (2,154) |  |  |
| Lesotho | 1,426 (3.8) | 54.0 (770) | 46.0 (656) | $F(3.60, 15,759) = 194.1$ | **<0.001** |
| **Wealth quintile** |  |  |  |  |  |
| Poorest | 9,048 (24.2) | 91.5 (8,275) | 8.5 (773) |  |  |
| Poorer | 7,386 (19.8) | 88.8 (6,557) | 11.2 (829) |  |  |
| Middle | 7,668 (20.5) | 85.3 (6,537) | 14.7 (1,131) |  |  |
| Richer | 7,437 (19.9) | 79.0 (5,873) | 21.0 (1,564) |  |  |
| Richest | 5,812 (15.6) | 61.5 (3,573) | 38.5 (2,239) | $F(3.77, 16,528) = 310.5$ | **<0.001** |
| **Maternal education** |  |  |  |  |  |
| No education | 10,299 (27.6) | 90.6 (9,330) | 9.4 (969) |  |  |
| Primary | 9,256 (24.8) | 85.9 (7,948) | 14.1 (1,308) |  |  |
| Secondary | 14,352 (38.4) | 79.8 (11,453) | 20.2 (2,899) |  |  |
| Higher | 3,444 (9.2) | 60.5 (2,084) | 39.5 (1,360) | F(2.96, 12,977) = 315.6 | **<0.001** |
| **Place of residence** |  |  |  |  |  |
| Urban | 12,929 (34.6) | 74.8 (9,673) | 25.2 (3,256) |  |  |
| Rural | 24,422 (65.4) | 86.6 (21,142) | 13.4 (3,280) | F(1, 4,382) = 297.0 | **<0.001** |
| **Maternal age group (years)** |  |  |  |  |  |
| 15–19 | 2,885 (7.7) | 85.6 (2,471) | 14.4 (414) |  |  |
| 20–24 | 9,379 (25.1) | 82.4 (7,728) | 17.6 (1,651) |  |  |
| 25–29 | 9,498 (25.4) | 81.0 (7,694) | 19.0 (1,804) |  |  |
| 30–34 | 7,572 (20.3) | 81.4 (6,166) | 18.6 (1,406) |  |  |
| 35–39 | 5,325 (14.3) | 83.6 (4,454) | 16.4 (871) |  |  |
| 40–44 | 2,214 (5.9) | 85.2 (1,886) | 14.8 (328) |  |  |
| 45–49 | 478 (1.3) | 87.0 (416) | 13.0 (62) | F(5.85, 25,646) = 6.2 | **<0.001** |
| **Parity** |  |  |  |  |  |
| Primipara (1st birth) | 8,456 (22.6) | 76.4 (6,460) | 23.6 (1,996) |  |  |
| 2nd–3rd birth | 13,381 (35.8) | 79.7 (10,669) | 20.3 (2,712) |  |  |
| 4th–5th birth | 8,221 (22.0) | 85.8 (7,050) | 14.2 (1,171) |  |  |
| 6th or higher birth | 7,293 (19.5) | 90.9 (6,636) | 9.0 (657) | F(2.91, 12,767) = 135.6 | **<0.001** |

**Note.** Values are unweighted observation counts with survey-weighted row percentages in parentheses. CoC = continuum of care; ANC = antenatal care; PNC = postnatal care. Complete CoC defined as adequate ANC ( $\geq 4$ visits with first-trimester initiation) + skilled birth attendance + postnatal care for the mother or newborn within 48 hours of birth. Incomplete CoC = failure to meet one or more of the three components. F-statistics are design-adjusted Pearson chi-square tests converted to F-statistics to account for the complex stratified cluster sampling design; reported at the level of the category variable. <sup>a</sup> Design degrees of freedom ( $df_2$ ) reflect the number of primary sampling units minus the number of strata in the pooled dataset. Bold p-values indicate statistical significance at $\alpha = 0.05$ . F-statistics for individual country rows are suppressed and reported at the category level.

### Neonatal mortality and its relationship with continuum of care

The overall pooled neonatal mortality rate (NMR) was 29.3 per 1,000 live births (95% CI: 27.6–31.0). Country-specific NMRs ranged from 20.4 per 1,000 in Kenya (95% CI: 17.7–23.0) to 41.0 per 1,000 in Nigeria (95% CI: 37.3–44.8), with Mali at 28.6, Congo DRC at 23.8, and Lesotho at 26.1 per 1,000 live births. Unadjusted comparison by CoC status revealed that births whose mothers received complete CoC had a lower NMR at 15.8 per 1,000 live births compared to 20.6 per 1,000 among those with incomplete CoC. Examination of the CoC score distribution revealed that births with zero care components had the lowest observed NMR (19.3 per 1,000), while births with one component had the highest (23.5 per 1,000), followed by two components (19.3 per 1,000) and three components (15.8 per 1,000). This non-monotonic pattern in the unadjusted analysis likely reflects selection into the zero-component group, comprising mothers in settings with effectively no access to formal healthcare infrastructure, a context associated with specific unmeasured mortality risk profiles rather than a causal protective effect of zero care.

### Association between complete CoC and neonatal mortality (Objective 2)

Table 4 presents the unadjusted and adjusted odds ratios (aOR) and adjusted prevalence ratios (aPR) from the survey-weighted logistic and log-binomial (Poisson) regression models. In the unadjusted analysis, complete CoC was associated with a 24% lower odds of neonatal death compared with incomplete CoC (cOR: 0.764, 95% CI: 0.583–1.000, p = 0.050). After adjustment for wealth, education, residence, parity, maternal age, child sex, and country, the association was attenuated and no longer statistically significant (aOR: 0.961, 95% CI: 0.717–1.289, p = 0.793; aPR: 0.962, 95% CI: 0.722–1.282, p = 0.792), indicating that the crude association was substantially confounded by sociodemographic factors and between-country differences in mortality levels. Among the adjustment covariates, maternal age demonstrated the most consistent and significant inverse associations with neonatal mortality. Compared with adolescent mothers aged 15 to 19 years, women aged 25 to 29 had the lowest adjusted odds of neonatal death (aOR: 0.503, 95% CI: 0.345–0.734, p < 0.001), followed by women aged 30 to 34 (aOR: 0.452, 95% CI: 0.294–0.697, p < 0.001). Female sex of the child was associated with significantly lower odds of neonatal death compared with male sex (aOR: 0.765, 95% CI: 0.636–0.920, p = 0.004). After accounting for individual covariates, substantial between-country differences in neonatal mortality persisted: compared with Nigeria (reference), all other countries showed significantly lower adjusted odds of neonatal death, with the strongest reductions in Lesotho (aOR: 0.340, 95% CI: 0.189–0.612, p < 0.001) and Kenya (aOR: 0.372, 95% CI: 0.279–0.495, p < 0.001).

**Table 4.** Survey-weighted logistic and Poisson regression results for the association between complete continuum of care and neonatal mortality.

| Variable | cOR | 95% CI | p | aOR<sup>a</sup> | 95% CI | p | aPR<sup>a</sup> (95% CI) |
| --- | --- | --- | --- | --- | --- | --- | --- |
| **Continuum of care (ref: Incomplete)** |  |  |  |  |  |  |  |
| Complete CoC | 0.764 | 0.583–1.000 | 0.050 | 0.961 | 0.717–1.289 | 0.793 | 0.962 (0.722–1.282) |
| **Wealth quintile (ref: Poorest)** |  |  |  |  |  |  |  |
| Poorer | - |  |  | 1.155 | 0.858–1.554 | 0.342 | 1.150 (0.862–1.535) |
| Middle | - |  |  | 1.079 | 0.803–1.451 | 0.614 | 1.077 (0.807–1.438) |
| Richer | - |  |  | 1.127 | 0.794–1.600 | 0.503 | 1.124 (0.798–1.581) |
| Richest | - |  |  | 1.152 | 0.746–1.779 | 0.524 | 1.147 (0.750–1.754) |
| **Maternal education (ref: No education)** |  |  |  |  |  |  |  |
| Primary | - |  |  | 1.042 | 0.794–1.368 | 0.767 | 1.042 (0.799–1.358) |
| Secondary | - |  |  | 0.876 | 0.662–1.158 | 0.353 | 0.880 (0.670–1.155) |
| Higher | - |  |  | 0.696 | 0.447–1.084 | 0.109 | 0.702 (0.455–1.083) |
| **Residence (ref: Urban)** |  |  |  |  |  |  |  |
| Rural | - |  |  | 1.123 | 0.881–1.431 | 0.349 | 1.120 (0.884–1.418) |
| **Parity (ref: Primipara)** |  |  |  |  |  |  |  |
| 2nd–3rd | - |  |  | 0.947 | 0.711–1.261 | 0.710 | 0.948 (0.717–1.254) |
| 4th–5th | - |  |  | 0.940 | 0.649–1.362 | 0.744 | 0.942 (0.656–1.353) |
| 6th+ | - |  |  | 1.166 | 0.761–1.786 | 0.481 | 1.162 (0.766–1.762) |
| **Maternal age (ref: 15–19 years)** |  |  |  |  |  |  |  |
| 20–24 | - |  |  | 0.606 | 0.427–0.859 | 0.005 | 0.615 (0.439–0.862) |
| 25–29 | - |  |  | 0.503 | 0.345–0.734 | <0.001 | 0.513 (0.355–0.740) |
| 30–34 | - |  |  | 0.452 | 0.294–0.697 | <0.001 | 0.462 (0.304–0.703) |
| 35–39 | - |  |  | 0.558 | 0.340–0.914 | 0.021 | 0.567 (0.351–0.917) |
| 40–44 | - |  |  | 0.603 | 0.341–1.067 | 0.083 | 0.612 (0.352–1.066) |
| 45–49 | - |  |  | 0.999 | 0.440–2.268 | 0.997 | 0.992 (0.453–2.175) |
| **Sex of child (ref: Male)** |  |  |  |  |  |  |  |
| Female | - |  |  | 0.765 | 0.636–0.920 | 0.004 | 0.770 (0.643–0.922) |
| **Country (ref: Nigeria)** |  |  |  |  |  |  |  |
| Mali | - |  |  | 0.501 | 0.381–0.659 | <0.001 | 0.510 (0.391–0.666) |
| Congo DRC | - |  |  | 0.575 | 0.442–0.746 | <0.001 | 0.583 (0.452–0.752) |
| Kenya | - |  |  | 0.372 | 0.279–0.495 | <0.001 | 0.380 (0.287–0.503) |
| Lesotho | - |  |  | 0.340 | 0.189–0.612 | <0.001 | 0.348 (0.195–0.620) |

**Note.** cOR = crude (unadjusted) odds ratio; aOR = adjusted odds ratio; aPR = adjusted prevalence ratio. Reference categories: incomplete CoC; wealth = poorest; maternal education = no education; residence = urban; parity = primipara; maternal age = 15–19 years; child sex = male; country = Nigeria. <sup>a</sup> Adjusted for all variables shown in the table simultaneously. Survey-weighted estimates accounting for stratification, clustering, and normalised pooled weights. All models use the Huber-White sandwich variance estimator for complex survey data.

### Dose-response analysis: CoC Score and neonatal mortality (Objective 2, continued)

Table 5 presents adjusted estimates for the categorical CoC score. Using zero components as the reference, each level of the CoC score was independently associated with higher adjusted odds of neonatal mortality compared with the reference group: one component (aOR: 1.753, 95% CI: 1.254–2.452, p = 0.001), two components (aOR: 1.701, 95% CI: 1.229–2.354, p = 0.001), and all three components (aOR: 1.576, 95% CI: 1.032–2.408, p = 0.035). This pattern, in which those receiving no components appear to have lower adjusted odds of neonatal death than those receiving partial or complete care, is consistent with selection into zero-component care: this group is predominantly composed of births in contexts of near-complete care absence, which in survival analyses of low-income settings can reflect a selected, lower-risk subpopulation rather than a causal protective effect of zero care. The p-value for the linear trend across the CoC score (treating score as continuous) was 0.085, indicating a marginally non-significant trend toward declining mortality with increasing CoC score when modelled continuously.

**Table 5.** Dose-response analysis: adjusted odds ratios and prevalence ratios for neonatal mortality by CoC score.

| CoC score (components received) | NMR<sup>a</sup> | aOR<sup>b</sup> | 95% CI | p | aPR<sup>b</sup> | 95% CI | p |
| --- | --- | --- | --- | --- | --- | --- | --- |
| 0 components - reference | 12.6 | 1.00 | - | - | 1.00 | - | - |
| 1 component | 21.8 | 1.753 | 1.254–2.452 | 0.001 | 1.726 | 1.246–2.392 | 0.001 |
| 2 components | 21.2 | 1.701 | 1.229–2.354 | 0.001 | 1.676 | 1.222–2.300 | 0.001 |
| 3 components (complete CoC) | 19.7 | 1.576 | 1.032–2.408 | 0.035 | 1.556 | 1.029–2.351 | 0.036 |
| p for linear trend (per unit increase in score) |  | 1.110 | 0.986–1.250 | 0.085 |  |  |  |

**Note.** NMR = neonatal mortality rate per 1,000 live births (unadjusted, survey-weighted). aOR and aPR adjusted for wealth quintile, maternal education, place of residence, parity, maternal age, sex of child, and country. Reference group: zero CoC components received. <sup>a</sup> Unadjusted weighted NMR per 1,000 live births. <sup>b</sup> All models adjusted simultaneously for all covariates listed in Table 3.

Survey-weighted marginal predictions (Figure 1) confirmed this pattern: the predicted probability of neonatal death was lowest in the zero-components group at 1.26% (95% CI: 0.91–1.61%), increased for one component (2.18%, 95% CI: 1.79–2.58%) and two components (2.12%, 95% CI: 1.82–2.41%), then declined among births with complete CoC (1.97%, 95% CI: 1.45–2.48%). Restricting to the comparison between incomplete and complete CoC (binary analysis), the predicted absolute risk difference was −0.07 percentage points (complete vs. incomplete CoC), equivalent to 0.74 fewer neonatal deaths per 1,000 live births attributed to complete CoC after full covariate adjustment.

**Figure 1.**
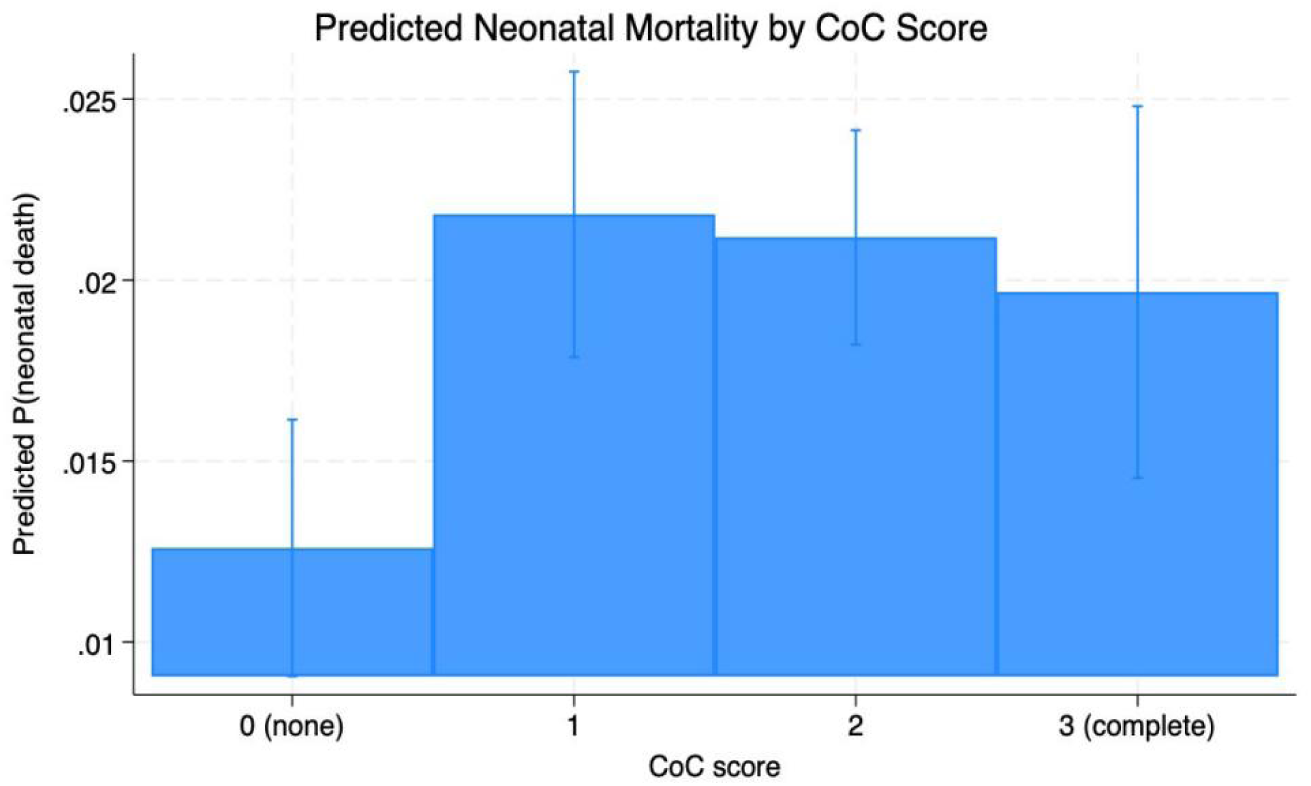
Marginal standardised predicted probabilities of neonatal death by continuum of care score. **Note.** Error bars represent 95% confidence intervals. Estimates are survey-weighted marginal predictions from the fully adjusted logistic model, standardised over the observed covariate distribution.

### Counterfactual analysis: Population attributable fraction (Objective 3)

G-computation was applied to the fully adjusted logistic regression model to estimate the Population Attributable Fraction (PAF) under the counterfactual scenario in which all women received complete CoC. The observed marginal predicted probability of neonatal death was 1.970% (19.70 per 1,000 live births). Under the counterfactual scenario of universal complete CoC, the predicted probability was 1.906% (19.06 per 1,000 live births), yielding an absolute risk reduction (ARR) of 0.064 percentage points and a number needed to treat (NNT) of approximately 1,562 mothers who would need to receive complete CoC to prevent one additional neonatal death. The overall PAF was 3.2%, indicating that approximately 3.2% of neonatal deaths in this pooled population could be prevented under a scenario of universal complete CoC coverage. The modest PAF reflects the low current prevalence of complete CoC (19.2%) and the attenuated adjusted association between complete CoC and neonatal mortality observed in the fully adjusted model. Country-specific PAF estimates ranged from approximately 2.1% in Mali to 8.4% in Kenya, where the country-specific protective association of complete CoC was strongest.

### Cross-country heterogeneity in the effect of CoC (Objective 4)

Table 6 present country-specific adjusted effect estimates and the results of the random-effects meta-analysis. Across the four countries with sufficient observations, the direction of the CoC effect was generally protective, though statistical significance varied markedly by country. The strongest and only statistically significant country-specific effect was observed in Kenya (aOR: 0.455, 95% CI: 0.256–0.810, p < 0.05), where complete CoC was associated with a 54.5% reduction in the adjusted odds of neonatal death. In contrast, Lesotho showed a non-significant association in the opposite direction (aOR: 1.447, 95% CI: 0.496–4.220), likely reflecting the small number of events (n = 19) in the neonatal death category and consequent imprecision. Congo DRC showed a substantial point estimate for a protective effect (aOR: 0.557, 95% CI: 0.175–1.772) that was non-significant due to wide confidence intervals. Mali similarly showed a non-significant trend toward protection (aOR: 0.885, 95% CI: 0.539–1.451). Nigeria was excluded from the country-specific meta-analysis due to the absence of complete-case observations in the meta-analysis subsample.

**Table 6.** Country-specific adjusted odds ratios and random-effects meta-analysis for the association between complete CoC and neonatal mortality.

| Country | n<sup>a</sup> | Events | aOR | 95% CI | Weight (%) | Pooled aOR<sup>b</sup> | Heterogeneity |
| --- | --- | --- | --- | --- | --- | --- | --- |
| Mali | 6,520 | 123 | 0.885 | 0.539–1.451 | 38.3 |  |  |
| Congo DRC | 9,717 | 186 | 0.557 | 0.175–1.772 | 13.3 |  |  |
| Kenya | 9,916 | 128 | 0.455 | 0.256–0.810 | 33.3 |  |  |
| Lesotho | 1,426 | 19 | 1.447 | 0.496–4.220 | 15.1 |  |  |
| **Overall (random-effects DL)** | **27,579** | **456** | **0.718** | **0.447–1.152** | **100.0** | **$z = -1.37$, $p = 0.170$** | **$I^2 = 38.9\%$, $Q p = 0.179$** |

**Note.** Country-specific aOR estimates from survey-weighted logistic regression models adjusted for wealth quintile, maternal education, place of residence, parity, maternal age, and sex of child. Nigeria was excluded from the meta-analysis due to insufficient complete-case observations in the cross-country subpopulation analysis. Pooled estimate derived using the DerSimonian-Laird random-effects method. DL = DerSimonian-Laird; $\tau^2$ = between-study variance. <sup>a</sup> Births within the 36-month PNC reference window with complete case data. <sup>b</sup> Random-effects pooled estimate.

The DerSimonian-Laird random-effects pooled estimate was aOR 0.718 (95% CI: 0.447–1.152, z = −1.37, p = 0.170), indicating a 28.2% non-significant reduction in the adjusted odds of neonatal mortality associated with complete CoC in the pooled meta-analysis. Moderate heterogeneity was detected (I² = 38.9%, 95% CI: 0.0–80.8%; Cochran Q = 4.91, df = 3, p = 0.179; τ² = 0.0885), which, while below conventional thresholds for high heterogeneity (I² ≥ 50%), suggests that between-country differences in healthcare system capacity, CoC measurement, and population risk profiles may moderate the relationship between CoC and neonatal mortality. The formal interaction test between country and CoC status in the pooled model was evaluated to formally quantify this heterogeneity.

## Discussion

This pooled analysis of 37,351 births across five sub-Saharan African countries found that complete continuity of maternal healthcare defined as adequate antenatal care with first-trimester initiation, skilled birth attendance, and postnatal care within 48 hours was received by fewer than one in five women (19.2%), and that this receipt ranged from 7.8% in Congo DRC to 47.8% in Lesotho. While complete CoC was associated with a 24% reduction in the crude odds of neonatal death, this association was substantially attenuated and rendered non-significant after simultaneous adjustment for wealth, education, residence, parity, maternal age, child sex, and country. The adjusted odds ratio of 0.961 (95% CI: 0.717–1.289) and the adjusted prevalence ratio of 0.962 (95% CI: 0.722–1.282) provide no evidence of an independent protective effect of complete CoC at the population level in this pooled sample. The population attributable fraction under a counterfactual scenario of universal complete CoC was 3.2%, reflecting both the low prevalence of complete CoC and the attenuated adjusted association. In the meta-analysis, the pooled random-effects estimate was directionally protective (aOR: 0.718) but non-significant, with moderate between-country heterogeneity (I² = 38.9%). Kenya was the only country in which complete CoC was associated with a statistically significant reduction in neonatal mortality (aOR: 0.455, 95% CI: 0.256– 0.810).

### Interpretation: Why the adjusted association was attenuated

The key finding of this analysis is not the adjusted null result but what it reveals about confounding in the CoC-mortality relationship. The attenuation from cOR 0.764 to aOR 0.961 after covariate adjustment is both statistically and substantively meaningful, with three principal interpretations. First, factors enabling women to access complete CoC - household wealth, higher education, urban residence, lower parity - independently associate with lower neonatal mortality risk through pathways beyond healthcare utilisation. Wealthier households have better nutrition, cleaner birth environments, greater access to emergency obstetric care, and higher capacity to respond to neonatal emergencies outside formal healthcare settings[39]. Higher maternal education is associated with superior birth preparedness, more effective breastfeeding, and timely care-seeking for neonatal danger signs[40]. These factors represent alternative pathways to neonatal survival that operate whether or not a woman engages with the formal care continuum. When adjusted for, the marginal contribution of the care continuum becomes smaller, as it no longer absorbs variance attributable to the broader socioeconomic environment.

Second, country fixed effects in the adjusted model absorb large baseline neonatal mortality differences between countries, partially correlated with CoC prevalence in the crude analysis. Nigeria, the largest country, had the highest NMR (41.0 per 1,000) and low CoC prevalence (15.0%). Countries with lower NMRs (Kenya: 20.4; Congo DRC: 23.8) had higher CoC prevalences. Conditioning on country, between-country mortality variance no longer drives the CoC estimate, and the within-country residual association is smaller and more precise. This pattern is consistent with literature on healthcare utilization and mortality in low-income settings: variation in care access and mortality is driven by structural determinants, and adjustment for these commonly attenuates associations[41]. Third, measurement imprecision in the PNC component may bias the CoC indicator toward the null. PNC within 48 hours is recalled in DHS surveys, and studies show systematic underreporting of early postnatal checks, especially for home births and high-parity women, which may misclassify some women who received care as not having received it[42]. This non-differential exposure misclassification tends to attenuate odds ratio estimates toward unity, potentially obscuring a true protective association.

### The dose-response paradox and its explanation

The dose-response analysis showed an unexpected pattern: marginal predicted probability of neonatal death was lowest in the zero-components group (1.26%) and higher for one, two, and three components before declining among those with complete CoC. This paradox is recognized in survival analyses from low-income, low-coverage settings[43]. Women receiving no formal care components are not randomly selected; they concentrate in areas with extreme geographic remoteness, community norms disfavoring formal healthcare, or nomadic populations with low health system contact. In these settings, neonatal mortality risk may be mediated by protective factors specific to the community context - experienced traditional birth attendants, extended family support, low exposure to hospital-acquired infection - rather than formal healthcare. Women engaging with one or two components but not the full continuum may represent those in transitional settings where incomplete formal healthcare has disrupted traditional practices without replacing them, creating a vulnerable group. This interpretation is supported by the non-significant linear trend (p = 0.085) and aligns with prior observations in West African and East African DHS analyses[43, 44].

### Cross-country heterogeneity and context-specificity

The moderate between-country heterogeneity in the meta-analysis (I² = 38.9%, Q p = 0.179) warrants attention, despite falling below significance thresholds. Kenya was the only country where complete CoC showed a significant protective effect on neonatal mortality (aOR: 0.455). This may be explained by Kenya’s investments in community health workers and the Linda Mama programme providing free maternity services since 2013, which improved maternal care coverage and quality[45]. The finding that CoC is most effective where service quality is highest aligns with the premise that the continuum of care framework operates through quality-sensitive care delivery rather than mere health system contact. Lesotho’s non-significant opposite estimate (aOR: 1.447) requires caution due to few neonatal events (n = 19) in the sample. With Lesotho’s high complete CoC prevalence (47.8%), detecting mortality benefits may require larger samples. Congo DRC’s wide confidence interval (0.175–1.772) reflects challenges in estimating country-specific effects where complete CoC prevalence is very low (7.8%), with a small, selected exposed group.

### Comparison with prior literature

The findings align with Alem and colleagues [17], who in analyzing 32 SSA countries using DHS-7 data found that complete CoC (defined as ANC4+, SBA, and at least one PNC visit) was received by a similarly low proportion of women (approximately 20%) and was significantly associated with lower neonatal mortality in crude but not fully adjusted analyses. This study extends their work by using DHS-8 data with more precise PNC measurement, applying a more rigorous ANC definition requiring first-trimester initiation with four or more visits, and quantifying population-level impact through formal G-computation. Kanyangarara and colleagues[18]reported that care continuum benefits depended heavily on care quality and healthcare system context, a finding replicated here through country-specific heterogeneity. The results also align with a multicounty study by Eshetu and colleagues[46], who found postnatal care indicators in DHS data were weakly associated with neonatal outcomes at the population level, attributing this to measurement limitations and genuine heterogeneity in PNC quality across settings.

### Strengths and limitations

The principal strengths of this analysis are its use of nationally representative, standardised DHS-8 data with explicit accounting for the complex sampling design; the large multi-country sample with diverse geographic and epidemiological contexts; the application of both logistic and Poisson regression to estimate odds ratios and prevalence ratios simultaneously; the formal counterfactual analysis; and the rigorous examination of cross-country heterogeneity through meta-analysis, interaction testing, and multilevel modelling. The use of DHS-8 data with the revised PNC measurement framework represents a methodological advance over prior pooled analyses.

Several limitations require acknowledgment. First, the cross-sectional design precludes causal inference: the association between CoC and neonatal mortality may have residual confounding by unmeasured variables including healthcare quality, household sanitation, maternal nutrition, and obstetric risk factors not in DHS. Country fixed effects and individual-level covariate adjustment mitigate but do not eliminate this concern. Second, PNC in DHS is measured by retrospective maternal recall, susceptible to recall and social desirability bias; misclassification likely trends toward underreporting, biasing the CoC effect toward null. Third, DHS does not capture content or quality during antenatal, delivery, or postnatal encounters, and healthcare contact does not guarantee evidence-based practices. Fourth, Nigeria was excluded from country-specific meta-analysis due to insufficient complete-case observations, limiting geographic representativeness. Fifth, the analytical sample was restricted to births within 36 months due to DHS-8 PNC reference window, potentially introducing a period effect if mortality trends differed in the earlier five-year window. Sixth, with five countries in the meta-analysis, power to detect heterogeneity and estimate random-effects mean is limited, and I² confidence intervals are wide.

### Policy implications

Although the adjusted analysis did not find a statistically significant independent association between complete CoC and neonatal mortality, the policy implications of the full set of findings are substantial and should not be interpreted as evidence that the continuum of care is ineffective. Rather, the results illuminate specific mechanisms and contextual conditions under which the framework does and does not translate into measurable survival benefit, and they point toward a more precise and differentiated policy agenda.

#### Address structural determinants alongside service delivery

The most important policy message from this analysis is that the protective effect of the maternal healthcare continuum is substantially mediated by and confounded with the broader socioeconomic determinants of neonatal health. Wealth quintile, maternal education, and place of residence were the strongest predictors of complete CoC receipt. Programmes that expand formal healthcare access without simultaneously addressing these structural barriers are unlikely to achieve equitable improvements in neonatal survival. Policies that increase household income and reduce financial barriers to care-seeking including free maternity service programmes, conditional cash transfers, and community-based health financing schemes are likely to have larger and more equitable effects on neonatal mortality than supply-side service delivery expansions alone [47]. The concentration of complete CoC in the richest quintile (38.5%) compared with the poorest (8.5%) represents a profound equity gap that demands targeted redistribution of healthcare resources toward underserved communities.

#### Prioritise care quality over contact frequency

Kenya’s significantly protective country-specific CoC effect, in the context of that country’s substantial investments in healthcare quality and community health worker programmes, supports the hypothesis that the CoC framework produces measurable neonatal mortality reductions in contexts where quality-assured care is delivered at each contact point. This implies that national programmes should shift emphasis from reporting contact rates the number of ANC visits attended or the proportion of facility deliveries toward monitoring the content and quality of care delivered within each contact. Signal function assessments, clinical audits of obstetric and newborn care practices, and expansion of emergency obstetric care at peripheral facilities are likely to amplify the survival benefit attributable to CoC utilisation.

#### Accelerate first-trimester antenatal care initiation

Only 30.9% of women across the five countries initiated antenatal care in the first trimester, which was the most restrictive bottleneck in the CoC cascade. Because first-trimester initiation enables early detection of anaemia, gestational hypertension, and infection, its importance extends beyond simple visit counts. Community-based awareness campaigns, integration of antenatal care registration into community health worker remit, and removal of user fees at first contact could each contribute to earlier initiation. Programmes targeting adolescent and primigravid women, who are at elevated risk of late presentation, deserve particular priority.

#### Expand postnatal care as the weakest link in the continuum

While overall PNC coverage within 48 hours was 64.8%, Congo DRC reported only 35.8%, representing the most critical gap in the care cascade for that country and suggesting that targeted investments in postpartum care are a priority. National postnatal care programmes should ensure that a standardised package of newborn care thermal protection, breastfeeding support, infection prevention, and danger sign recognition is consistently delivered at discharge and through a first community contact within 24 to 48 hours. The DHS-8 finding that 40.5% of newborns in the pooled sample had no postnatal check by a health provider within two days of birth represents a substantial missed opportunity for the prevention of infection-related neonatal deaths, which account for approximately one-third of all neonatal mortality in SSA [3].

#### Tailor interventions to the country context

The moderate cross-country heterogeneity in the CoC-mortality relationship underscores that no single programme design will be optimal across all settings. Congo DRC, with a complete CoC prevalence of 7.8% and a high neonatal mortality rate, presents a fundamentally different programmatic challenge from Lesotho, where nearly half of women already receive complete CoC and the marginal benefit of further coverage expansion is likely smaller than improving care quality for those already engaged. National health ministries and development partners should use country-specific DHS-8 data to identify the specific bottleneck in the care cascade whether at the ANC, delivery, or postnatal stage and to design responsive interventions that address that specific stage most efficiently.

#### Invest in data systems that capture care quality

The limitations of DHS-based CoC analysis highlight the need for longitudinal, linked facility-level data that can track individual women through the full care pathway and document the content of care at each contact. The absence of quality indicators in the DHS means that a woman who attended four antenatal visits with no blood pressure measurement or malaria prophylaxis is classified identically to one who received the full recommended package of care. Facility-based administrative data linked to vital registration, where feasible, would substantially improve the evidence base for the continuum of care framework in SSA and enable more precise identification of the specific elements of care that drive neonatal survival benefit.

### Conclusions

This pooled analysis of DHS-8 data from five sub-Saharan African countries shows that complete continuity of maternal healthcare remains underutilised, with only 19.2% of women receiving adequate antenatal care, skilled birth attendance, and postnatal care within 48 hours. While unadjusted data suggest a protective association with neonatal mortality, this association is explained by sociodemographic determinants of both care access and newborn survival rather than an independent causal effect. The null adjusted finding should not be misread as evidence that maternal healthcare does not matter for neonatal survival; it indicates that in SSA’s current epidemiological context, care utilisation and quality are so unevenly distributed that the benefit of completing the formal care pathway is difficult to detect above structural inequality. Achieving the SDG 3.2 neonatal mortality target in SSA requires simultaneous action on quality healthcare service supply, the barriers determining access, and factors that mediate the translation of care contact into survival benefit. The significant CoC effect observed in Kenya provides proof that the CoC framework can deliver measurable neonatal mortality reductions when embedded in a system delivering quality-assured care, offering a model for other regional countries to emulate.

## Acknowlegements

We thank BANGA for their scholarly support. We acknowledge the support of the UTG, UG, and UFL for their institutional support.

## Author contributions

SC and AB conducted all statistical analyses, drafted the original manuscript including the background, methods, results, and discussions sections. DD conceptualized the study. PT reviewed the manuscript.

## Data availability

This data is open access and is available to registered users of the https://www.dhs.org/database

## Declarations

### Consent for publication

This study used publicly available secondary, deindentified, and aggregate data. Consent for publication is not applicable.

